# Beyond latent and active – a scoping review of conceptual frameworks and diagnostic criteria for tuberculosis

**DOI:** 10.1101/2023.07.05.23292171

**Authors:** Syed MA Zaidi, Anna K Coussens, James A Seddon, Tamara Kredo, Digby Warner, Rein M G J Houben, Hanif Esmail

**Author notes:** Corresponding Author: Syed MA Zaidi.

## Abstract

**Background:** There is growing recognition that tuberculosis (TB) infection and disease exists as a spectrum of states beyond the current binary classification of latent and active TB. Our aim was to systematically map and synthesize published conceptual frameworks for different TB states from the literature.

**Methods:** We searched MEDLINE, Embase and EMcare for systematic and narrative reviews without date restrictions. We included articles that explicitly described greater than two states for TB. We conducted a thematic and frequency analysis for terminologies, conceptual definitions and diagnostic criteria for defined TB states.

**Results:** We identified 37 articles that met our inclusion criteria. All included articles were published after 2009. We identified eight broad conceptual themes that were used to categorize TB states and to calculate their frequency among included articles. These states were: State 0: *Mycobacterium tuberculosis* (*Mtb*) elimination by innate immune response (n=23/37, 62%); State I: *Mtb* elimination by acquired immune response (n=28/37, 76%); State II: *Mtb* infection not eliminated but controlled by immune system (n=34/37, 92%); State III: *Mtb* infection not controlled by the immune system (n=21/37, 57%); State IV: bacteriologically positive without symptoms (n=23/37, 62%); State V: signs or symptoms associated with TB (n=36/37, 97%); State VI: severe or disseminated TB disease (n=11/37, 30%); and State VII: previous history of TB (n=5/37, 14%). We found 27 additional variations within these themes that were labelled as “sub-states.” Articles varied in the terminology used to describe conceptual states and similar terms were often used to describe different concepts. Diagnostic criteria were provided in 27 articles and were also applied inconsistently.

**Conclusion:** Terminologies and definitions for TB states are highly inconsistent in the literature. Consensus on a framework that includes additional TB states is required to standardize communication in scientific publications as well as to inform advancements in research, clinical and public health practice.

**Panel: Research in context:** *Evidence before this study:* The current paradigm of tuberculosis (TB) is based on a binary classification into “latent” infection and “active” disease states. In recent years, there has been growing recognition that this binary classification does not accurately reflect the complex pathophysiology of the disease process and that it may also be inadequate for informing research and programmatic advances for global TB elimination. While a number of articles have proposed multiple states of infection and disease, no previous study has mapped and synthesized evidence from published literature to inform an overarching and inclusive staging framework. We conducted a comprehensive search on MEDLINE, Embase and EMCare databases for systematic or narrative review articles or commentaries with terms related to TB and “states”, “stages,” “paradigm” “framework” or “spectrum” without date restrictions. We included 37 articles that explicitly described TB as a multi-state, i.e., beyond latent and active disease.

*Added value of this study:* To our knowledge, this is the first study to systematically review conceptual frameworks, terminologies and diagnostic criteria for TB states beyond the latent and active paradigm. We identified that there is substantial variation in the number of TB states described in the literature, as well as in the concepts used to categorize them. Terms used for describing TB states and their diagnostic criteria were also inconsistently applied.

*Implications of all the available evidence:* Our review highlights the need for a clear consensus on the overall conceptual framework, terminology and diagnostic criteria for TB states. The inconsistency in TB states among articles included in our review reflects diverse perspectives, academic interests and research priorities. The consensus process should therefore aim to be inclusive so that a proposed framework can be acceptable to a broad range of stakeholders including clinicians, researchers, public health and policy practitioners, as well as to individuals living with or with experience of TB.

## Introduction

An estimated 10.6 million individuals fell ill with tuberculosis (TB) in 2021 and the disease caused 1.6 million deaths globally (1). TB elimination priorities, diagnostic approaches and treatments for the past several decades have been based on a binary understanding of the disease, described as having either latent infection or active (clinical) disease (2). While this simplicity has facilitated the development of programmatic management of TB since it was introduced over three decades ago, it has several limitations. Recent understanding of TB has revealed a more complex pathophysiology with a spectrum of disease states, ranging from the time of exposure to *Mycobacterium tuberculosis* (*Mtb*) bacilli to symptomatic, “active” disease (3). Individuals within this spectrum likely differ in their prognostic and treatment outcomes, however, their clinical management continues to be based on a binary understanding of the disease (4). Individuals with early and unrecognized TB states may also be an important source of community transmission and are therefore critical from an epidemiological perspective (5). From a programmatic viewpoint, individuals with early disease can be potentially misdiagnosed or missed by the health system. This is becoming increasingly relevant as active case-finding interventions are being scaled-up across high-burden countries (6).

Numerous articles have been published recently motivating that TB should be considered as a spectrum of disease states. However, authors often take different perspectives (e.g. immunological, microbiological, public health) and it is currently unclear which states have been commonly described in literature, what terms have been used to describe them, how should they be defined, as well as what diagnostic tests can be used to identify them. There is a need to map TB states beyond latent and active disease described in the literature to help guide researchers, clinicians and policymakers. Therefore, the aim of this scoping review was to collate the terminology, conceptual and diagnostic definitions utilized to describe the spectrum of TB states beyond the latent and active paradigm.

## Methods

The scoping review was conducted based on a protocol that was registered at the Open Science Framework (OSF) (7). The review consisted of the following research questions: 1*) what novel conceptual states have been described for TB beyond latent and active disease and how can they be categorized; 2) what nomenclature has been used to describe novel TB states; 3) what are the diagnostic definitions for novel TB states; 4) which conceptual states of TB have been described as potentially infectious and as representing TB disease; and 5) how have the concepts and terms used for TB states have changed over time.* Due to the lack of working definitions and absence of conceptual clarity on TB disease states, a scoping review methodology was considered appropriate. The protocol was developed in accordance with the recommendations provided by the Joanna Briggs Methods Manual for Scoping Reviews (8). The scoping review objectives were developed with input from all authors. The inclusion criteria and charting methods were developed by one author (SMAZ) and reviewed by another author (HE) to improve the validity. The study protocol was developed by two authors (SMAZ and HE) and reviewed by the remaining authors. The Preferred Reporting Items for Systematic reviews and Meta-Analyses Extension for Scoping Reviews (PRISMA-ScR) was utilized to guide the reporting (9).

### Search Strategy and Data Sources

To address research questions 1-4, we conducted a systematic search on MEDLINE, Embase and EMcare databases on Ovid for terms related to TB states, stages or spectrum. The search was conducted in December 2022 and no date restrictions were applied. The complete list of terms utilized in the scoping review are described in Supplementary Table 1. A filter for “Review Articles” was applied in Ovid. The search strategy was developed with input from a specialist librarian at the UCL Great Ormond Street Institute of Child Health Library. In addition to the database search for published review articles, to address research question 5, we separately charted the history of classification of TB used by the National TB Association (NTA) of the United States of America, from its foundation in 1904 to the current day. Transactions of the annual meetings of the NTA and the published Diagnostic Standards (in there various formats) from 1917-2016 were obtained (10). The findings from this historical review are summarized in Box 1.

#### Box 1: TB Disease states: historical perspective

The articles that form part of this scoping review are all recent which in part reflects the search strategy, selection of databases and inclusion criteria. However, discussions and debates on the classification of TB have been ongoing since well before Koch’s discovery of *Mycobacterium tuberculosis*. Historical literature and the Diagnostic Standards and Classification of Tuberculosis of the (American) National Tuberculosis Association (published periodically since 1917) highlight an evolving classification of TB driven by changing priorities for TB control, available diagnostics and treatments (10).

As early as the early nineteenth century, authors referred to and discussed the merits of staging systems for TB with terminology such as *phthisis incipiens* (incipient TB), *phthisis confirmata* (confirmed TB) and *phthisis desperata* (desperate/severe TB) in use (46, 47). By the early twentieth century there were numerous staging systems reflecting different perspectives, for instance based on pathology, diagnosis, extent, presentation or disease course (47–49). It was recognized that there was a tension between complex classification systems capturing multiple dimensions of the disease accurately vs. simplistic approaches such as the 3-stage Turban-Gerhardt classification based on disease extent that was could be more easily applied and hence widely implemented but proved unsatisfactory (48, 49).

While TB continued to be largely managed in sanatoria, microbiological, radiological and other diagnostic tools to assess the state of disease were becoming increasingly available and the need for a standardized approach to disease classification was recognized to aid statistical comparisons and clinical communication. In particular, it was considered important to have a consistent approach to classifying the condition of the patient at entry into the sanatoria (reflecting prognosis) and the condition during follow-up and at discharge (reflecting the degree of cure/healing and likelihood of relapse). Soon after inception of the National Tuberculosis Association in 1904, a *Committee on Clinical Nomenclature* was formed (50, 51). Discussions, surveys and consensus-building for over a decade led to the development of the 1^st^ Diagnostic Standards and Classification of Tuberculosis in 1917 (52). Here the classification of pulmonary TB in adults consisted of 3 dimensions each clearly defined; (i) the extent of pulmonary lesions – initially categorized as incipient, moderately advanced and far advanced (ii) symptoms – categorized slight/none, moderate (iii) clinical/treatment status – initially suspicious case, definite case, arrested/apparently arrested case. Over subsequent iterations details were added, clinical status became more nuanced, bacteriology was increasingly emphasized and there were changes to terminology. However, the broad classification approach remained similar with a focus on stratification by disease extent and clinical status (51).

The availability of antibiotics from the 1950’s for both curative and preventive therapy resulted in a radical change in the approach to managing tuberculosis and eventually led to a shift in disease classification (53). Prognostic considerations and clinical status became less relevant as treatment remained the same for all individuals and prolonged follow-up for relapse was not required. The 13^th^ edition of the Diagnostic Standards in 1974 were a major departure from previous classifications highlighting 5 states; *0: No tuberculosis exposure, not infected, I: Tuberculosis exposure, not infected, II: Tuberculosis infection, without disease, III: Tuberculosis infection, with disease, IV: Tuberculosis suspect* (53). Central to this classification was diagnosis of infection requiring evidence of immune sensitization without evidence of disease (symptoms, radiological or microbiological) and of disease through microbiological confirmation. Finally in 1999, the term *latent* was prefaced to *tuberculosis infection, no disease* to distinguish from *clinically active tuberculosis*, leading to the binary paradigm in use to the present day (54).

### Inclusion Criteria

Eligible articles explicitly described TB as a multi-state disease, i.e. provided description of states beyond the existing two-state latent and active disease paradigm. Included articles consisted of reviews pertaining to a range of topics including TB pathogenesis, symptomatology, immunology and bacteriology, diagnostics, vaccinology and transmission. Only articles published in the English language were eligible for inclusion. Articles describing specific diagnostic approaches or biomarkers without describing distinct conceptual states were excluded. Articles limited to descriptions of TB pathology or bacteriology without describing TB conceptual states within individuals were excluded. Articles with a focus on childhood or extra-pulmonary TB only were excluded.

### Screening, Data Abstraction and Charting

Outputs from the Ovid searches from the three databases were uploaded on Covidence and duplicates were removed (11). Two authors (SMAZ and HE) independently reviewed abstracts for inclusion. Conflicts were resolved through discussions based on the inclusion and exclusion criteria. Full-text articles were assessed for eligibility by one author (SMAZ) and reviewed by a second (HE). Conflicts on inclusion of articles after full-text reviews were resolved through review by a third author (RMGJH).

The data-charting table was developed on MS Excel. Summary characteristics of the included articles, textual descriptions of TB states, concepts and terminologies were abstracted. Conceptual descriptions of TB states described as part of diagrams or within texts for each article were tabulated. Due to the variation in frequency, descriptions and terminologies utilized for disease states between articles, textual descriptions with common concepts were thematically derived and coded. These conceptual themes were labelled as distinct “*states*” and were subsequently categorized numerically. Extended descriptions or minor variations within themes were separately coded in order to capture all described states. These were categorized as “*sub-states*” and categorized alphanumerically. Frequency of each state and sub-state among included articles were calculated. Two authors (SMAZ and HE) developed the coding and all authors reviewed the development of the final descriptions of TB states. Data-abstraction and coding was conducted by two authors (SMAZ and HE) and conflicts on the coding were resolved by mutual discussion. Disagreements on coding for TB states were resolved by a third author (RMGJH). One author (HE) reviewed proceedings from the NTA and abstracted historical timelines for the usage of various terminologies for describing TB disease states To record diagnostic criteria, outcomes of diagnostic tests and symptomatology described for each state were extracted from the articles. These were categorized as positive, positive/negative (i.e. test could be either positive or negative), negative, or not described. Frequency statistics describing diagnostic tests and symptomatology as positive or positive/negative for each state were calculated. Frequency of articles describing each state as potentially infectious and as representing TB disease were also calculated.

## Results

### Literature search

A total of 2,138 articles were identified through the database search (Figure 1). A further seven articles were identified through recommendations of the contributing authors. After removing duplicates, 1,512 article abstracts were screened, from which 64 full-text articles were assessed for eligibility. A total of 37 articles were included in the final qualitative synthesis. Most excluded articles (89%, n=24/27) did not describe TB as a multi-state disease. Articles focused on a singular aspect of the disease, such as pathology without describing states that would be of clinical or epidemiological relevance, were also excluded (11%, n=3/27). Nearly all of the included articles had conceptual diagrams with descriptions outlining disease states and their transitions.

**Figure 1:**
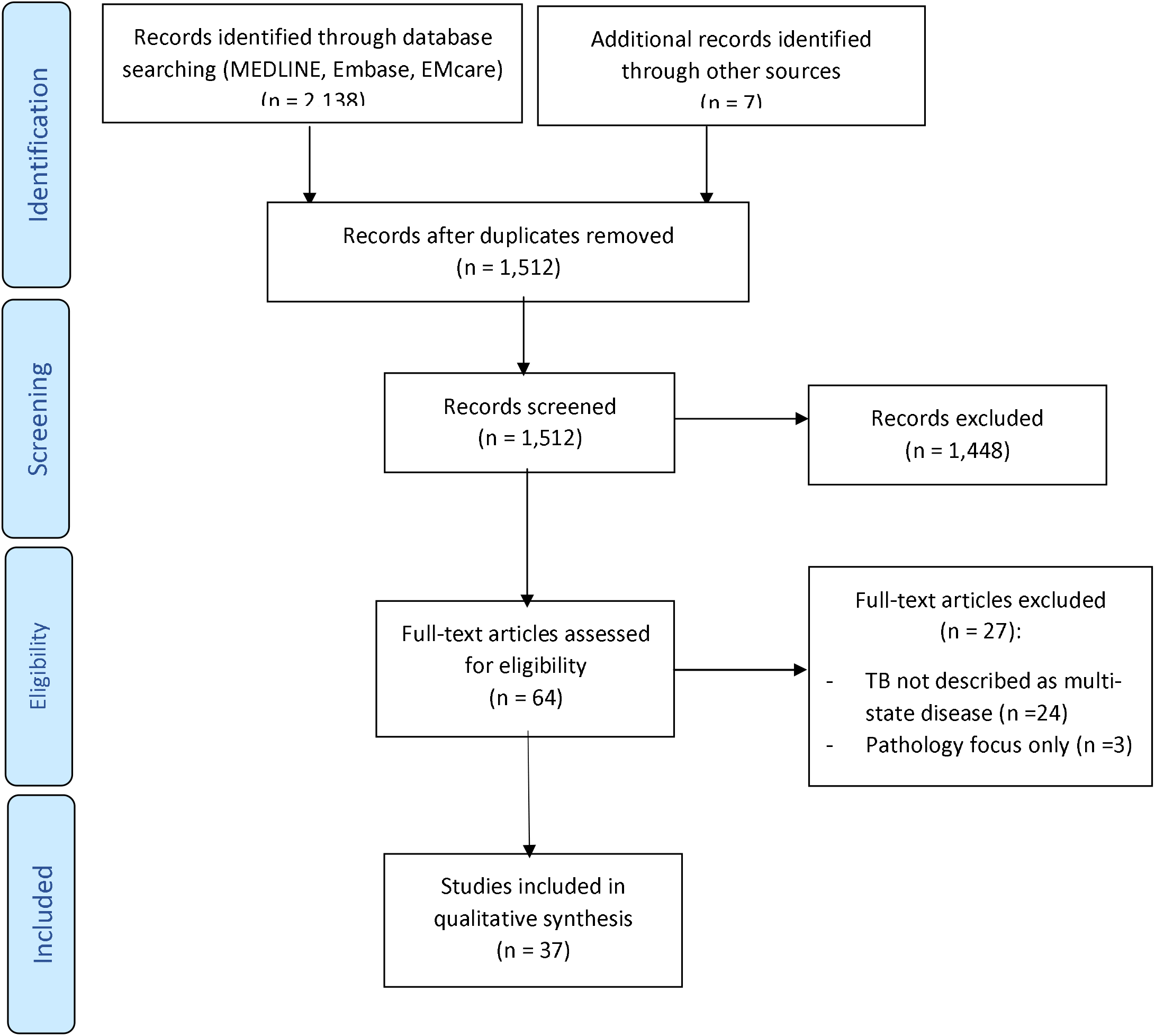
Selection of sources of evidence

### Characteristics of the included articles

Most articles consisted of narrative reviews (78%, n=29/37) (Table 1). Three articles (8%) were perspectives, two were systematic reviews and one a mathematical model. Most articles were primarily focused on novel understandings of TB pathogenesis (46%, n=17/37). The remaining articles described novel diagnostic approaches for TB (27%, n=10/37), advances in vaccine development (8%, n=3/37), epidemiological or transmission implications of early TB states (16%, n=6/37) and radiological findings of early TB (3% n=1/37). Most articles (91%, n=34/37) were written by primary authors based in high-income countries.

**Table 1:** Characteristics of the articles included in the scoping review

| **Reference** | **Year** | **Title** | **Article Aims & Objectives** | **Type of Article** |
| --- | --- | --- | --- | --- |
| Barry 3rd et al.(3) | 2009 | The spectrum of latent tuberculosis: rethinking the biology and intervention strategies | Review of biology of latent TB, host responses and drug development | Review article |
| Young et al.(13) | 2009 | Eliminating latent tuberculosis | Review of heterogeneity in TB physiology and host immune response and impact on therapeutic and diagnostic approaches | Review article |
| Chao and Rubin(14) | 2010 | Letting sleeping dogs lie: does dormancy play a role in tuberculosis? | Description of in vitro models of TB dormancy and resuscitation during infection. | Review article |
| Achkar and Jenny-Avital(15) | 2011 | Incipient and Subclinical Tuberculosis: Defining Early Disease States in the Context of Host Immune Response | Review of early TB disease states and their impact on biomarker, diagnostics and drug discovery | Review article |
| Gideon and Flynn(16) | 2011 | Latent tuberculosis: what the host “sees”? | Review of human and non-human primate models of tuberculosis that have led to new concepts of latent tuberculosis | Review article |
| Kunnath-Velayudhan et al.(17) | 2011 | Immunodiagnosis of Tuberculosis: a Dynamic View of Biomarker Discovery | Review of clinical spectrum of M. tuberculosis infection, evolution of granulomatous lesions and granuloma structural changes | Review article |
| Lawn et al.(18) | 2011 | Changing Concepts of “Latent Tuberculosis Infection” in Patients Living with HIV Infection | Review of spectrum of TB bacteriology and immunology and implications for HIV associated TB | Review article |
| Sridhar et al.(19) | 2011 | Redefining latent tuberculosis | Review of latent TB biology, immunology and diagnostics | Review article |
| Esmail et al.(20) | 2012 | Understanding latent tuberculosis: the key to improved diagnostic and novel treatment strategies | Review of current diagnosis and treatment of latent Mtb infection and developments in understanding the biology of latency | Review article |
| Joel D. Ernst(21) | 2012 | The immunological life cycle of tuberculosis | Descriptive framework for investigating immunity to M. tuberculosis to guide vaccine discovery and development. | Review article |
| Delogu et al.(22) | 2013 | The biology of mycobacterium tuberculosis infection | Review of advances in TB biology and pathogenesis and evolving paradigm of latent Mtb infection | Review article |
| Dowdy et al.(23) | 2013 | Is Passive Diagnosis Enough? The Impact of Subclinical Disease on | Estimate population-level impact of TB case-finding strategies in the presence of subclinical disease. | Mathematical Epidemic Model |
|  |  | Diagnostic Strategies for Tuberculosis |  |  |
| Delogu and Goletti(24) | 2014 | The Spectrum of Tuberculosis Infection: New Perspectives in the Era of Biologics | Review of TB disease spectrum | Review article |
| Esmail et al.(25) | 2014 | The ongoing challenge of latent tuberculosis | Review of current understanding of latent TB and highlight novel diagnostic, drug regimens and vaccines development approaches | Review article |
| Nunes-Alves et al.(26) | 2014 | In search of a new paradigm for protective immunity to TB | Review of advances in immune control of *M. tuberculosis* and implications for vaccine design and evaluation | Review article |
| Salgame et al.(27) | 2015 | Latent tuberculosis infection – Revisiting and revising concepts | Host, clinical and epidemiologic features of latent *Mtb* infection and diagnostic development | Review article |
| Pai and Behr(28) | 2016 | Latent *Mycobacterium tuberculosis* Infection and Interferon-Gamma Release Assays | Review of tests for latent TB diagnosis | Review article |
| Pai et al.(29) | 2016 | Tuberculosis | Review of the global TB epidemic | Review article |
| Petrucchioli et al.(30) | 2016 | Correlates of tuberculosis risk: predictive biomarkers for progression to active tuberculosis | Review of biomarkers for progression to active TB | Review article |
| Cadena et al.(31) | 2017 | Heterogeneity in tuberculosis | Review of TB biology and heterogeneity observed in the human disease | Review article |
| Drain et al.(32) | 2018 | Incipient and Subclinical Tuberculosis: a Clinical Review of Early Stages and Progression of Infection | Review of current understanding of TB pathogenesis, immunology, clinical epidemiology, diagnosis, treatment, and prevention | Review article |
| Kik et al.(33) | 2018 | An evaluation framework for new tests that predict progression from tuberculosis infection to clinical disease | Descriptive framework for generating evidence to inform the development of policy guidance for tests for incipient TB | Review article |
| Lin et al.(34) | 2018 | The End of the Binary Era: Revisiting the Spectrum of Tuberculosis | Review of data supporting the spectrum of *M. tuberculosis* infection in humans and nonhuman primates | Review article |
| Sousaa and Saraiva(35) | 2018 | Paradigm changing evidence that alter tuberculosis perception and detection: Focus on latency | Review of new understandings of TB latency to inform research and elimination | Review article |
| Houben et al.(36) | 2019 | Spotting the old foe—revisiting the case definition for TB | Novel disease case definitions for TB | Perspective |
| Lewinsohn and Lewinsohn (37) | 2019 | New Concepts in Tuberculosis Host Defense | Review of TB immunology and novel host-directed therapies | Review article |
| McHenry et al.(38) | 2020 | Genetics and evolution of tuberculosis pathogenesis: New perspectives and approaches | Descriptive framework on possible transitions from pathogen exposure to TB disease and associated genetics studies | Review article |
| Boom et al.(39) | 2021 | The knowns and unknowns of latent *Mycobacterium tuberculosis* infection | Review of immune responses to Mtb in humans and nonhuman primates, new concepts and major knowledge gaps in understanding of latent Mtb infection | Review article |
| Kendall et al.(40) | 2021 | The Epidemiological Importance of Subclinical Tuberculosis A Critical Reappraisal | Conceptual framework for subclinical TB and transmission | Perspective |
| Migliori et al.(41) | 2021 | The definition of tuberculosis infection based on the spectrum of tuberculosis disease | Review of TB as a disease spectrum | Review article |
| Alebouyeh et al.(42) | 2022 | Feasibility of novel approaches to detect viable *Mycobacterium tuberculosis* within the spectrum of the tuberculosis disease | Review of feasibility of new approaches to detect TB bacilli across the full spectrum disease | Review article |
| Chandra et al.(43) | 2022 | Immune evasion and provocation by *Mycobacterium tuberculosis* | Review of TB biology, immunity and development of host-directed therapies, biomarkers and vaccines. | Review article |
| Esmail et al.(2) | 2022 | Mind the gap - Managing tuberculosis across the disease spectrum. | Review of tuberculosis as a spectrum of disease states, new diagnostic approaches and clinical trials | Review article |
| Houben et al.(5) | 2022 | Tuberculosis prevalence: beyond the tip of the iceberg | Novel stages and disease thresholds for TB | Perspective |
| Scriba et al.(44) | 2022 | Challenges in TB research | Challenges in developing new diagnostics and vaccines for TB control | Perspective |
| Yoon et al.(45) | 2022 | CT and 18F-FDG PET abnormalities in contacts with recent tuberculosis infections but negative chest X-ray | Systematic review of CT and PET-CT imaging of asymptomatic TB contacts with normal X-rays | Systematic review |
| Sossen et al.(4) | 2023 | The Natural History of Untreated Pulmonary Tuberculosis in Adults: A Systematic Review and Meta-Analysis | Quantify progression and regression across the spectrum of TB disease of individuals with untreated TB during the pre-chemotherapy era | Systematic Review |

All the articles were published from 2009 onwards, with 13 (35%) being published during or after 2020. This reflects the relative novel conceptualization of TB as a multi-state disease within contemporary literature. However, as described in Box 1, such conceptual states were recognized and described in the pre-antibiotic era.

### Conceptual definitions of TB states

A total of eight distinct conceptual themes were identified that were categorized as TB states (Table 2). We categorized the first state as **State 0** and this was described in 23 (62%) articles. The conceptual theme for this state was elimination of *Mtb* through innate immunity due to absence of detectable acquired immune memory to *Mtb* infection. Uninfected individuals were also categorized as State 0. The most common terms for this state were ‘*innate immune response”, “innate immune elimination”* and *“uninfected.”* **State I** described in 28 (76%) articles was conceptually similar to State 0 in that the infection was eliminated. However, the elimination was described as involving an acquired immune response. This host response was frequently described as detectable and these individuals were categorized as distinct from those who were uninfected. The most used terms for this state were “*acquired immune,” “adaptive immune response*” and “*latent tuberculosis infection (LTBI).*” Individuals in **State II** described in 34 (92%) articles were categorized as those in whom *Mtb* has not been eliminated, however, the infection was described to be controlled by the immune system. This state was most commonly described as “*quiescent infection*”, “*latent tuberculosis infection*” and “*incipient TB*”. **State III** described in 21 (57%) articles was categorized as individuals in whom there was a failure to control the infection or a breakdown of immune control. The most commonly used terms for this state were *“incipient”, “incipient TB infection”,* and *“incipient TB disease.”* **State IV** described in 23 (62%) articles categorized individuals that were bacteriologically positive for TB but had no symptoms, or had symptoms that were not recognized or were not seeking care for symptoms. The state was most frequently described as “subclinical” or “subclinical disease”. In **State V,** individuals had TB symptoms and signs and were actively seeking care. This state was described in 36 (97%) of articles. This state was described as *“active TB disease”, “clinical TB”* or as *“disease.”* **State VI** described in 11 (30%) articles was differentiated from the previous state through terms describing increased disease severity such as cavitary disease, disseminated or extensive disease. The most frequently utilized term for this state was “*Cavitary/Disseminated*”. **State VII** grouped all individuals with a previous history of TB including those previously diagnosed, treated, or cured. This state was least frequently described, featuring in only 5 articles (14%). Articles varied in the use of conceptual themes to describe the TB spectrum (Table 3).

**Table 2:** TB states conceptual definitions and terminology

| State | Conceptual Theme | Terminology Used to Describe State | Articles Describing State (n) | % |
| --- | --- | --- | --- | --- |
| **State 0** | Infection has been eliminated by innate immune response | **Uninfected, Innate immune Response, Innate immune elimination**, No Disease (TB Negative), Eliminated TB infection, Infection controlled/ eliminated / Stable, Uninfected individual, No M. tuberculosis infection, Cleared, Latent Clearance, Uninfected, Initial infection, TB infection, Innate Immune Cell Recruitment, Colonization and Early Clearance, Infection, Resistance to Infection, Stage 1, Elimination of infection (self-resolving infection), Infection eliminated (with innate immune) | 23 | 62% |
| **State I** | Infection has been eliminated by acquired immune response | **Acquired Immune, Adaptive Immune Response, LTBI**, No Disease (Latent TB), Acquired Immune Response, Acquired Immune Elimination – memory, LTBI – Asymptomatic, Eliminated TB infection, Acquired Immune, Infection controlled/ Eliminated / Stable, Non-progression or Self-cure from Subclinical Stage, Primary or secondary M. tuberculosis infection, LTBI – Reverter, Latent Clearance, Reduced Reactivation Risk, Adaptive Immune Phase, Transient Infection, Adaptive Immune Cell Activation, Resister, Clearance of Infection, Mtb infection, Latent TB infection, Asymptomatic - LTBI – Incident | 28 | 76% |
| **State II** | Infection is not eliminated but is controlled by the immune system | **Quiescent Infection, Latent Mtb infection, LTBI, Incipient TB**, Latent Infection, No Disease (Latent TB), Quiescent, Unstable LTBI, Latent Non-infectious tuberculosis, Mtb infection, Immunological equilibrium (latency), Latent Subclinical or Low Grade TB, Primary Infection, Latent Infection - Dormant / Persistent, Traditional LTBI, Incipient TB, Caseating granuloma, Persistent TB, Containment of infection (Established Latency) | 34 | 92% |
| **State III** | Infection is not controlled by the immune system | **Incipient, Incipient TB infection, Incipient TB disease**, No Disease (Latent TB), Active Infection, Subclinical - TB Disease, Asymptomatic, Incipient disease, Unstable LTBI Incipient, Infectious Tuberculosis, Active Severe TB, Resuscitation, Reactivation TB, Percolating Infection, Progressor / Incipient TB, Stage 2, Failure to Contain Infection (subclinical active TB), Culture-Negative TB | 21 | 57% |
| **State IV** | Individual is bacteriologically positive but has no symptoms or clinical features | **Subclinical, Subclinical TB Disease, Asymptomatic,** Bacteriologically Positive, Subclinical Active Phase, Subclinical - No Symptoms, Additional Infectious Tuberculosis, Subclinical TB with no Signs/Symptoms, Subclinical Disease, Subclinical / Incipient, Active Infection, Incipient TB, Asymptomatic - Pre-clinical, Stage 3, Failure to contain infection (Primary Active TB), Subclinical Infection, Quiescent | 23 | 62% |
| **State V** | Individual has signs or symptoms associated with TB | **Active TB Disease, Clinical TB, Disease (Active TB), Disease,** Minimal, Active TB Disease - Symptomatic Clinical Bacteriologically Negative, Pre-diagnostic TB Disease, Clinical TB, Subclinical - Symptoms not Recognized, Clinical Disease (Symptoms) - Infectious Period, Active Clinical TB Disease, Chronic TB, Reactivation / Secondary TB, Reactivation, TB Disease, Transmission, Active TB, Smear Negative, Active TB - Caseating Granuloma, At Risk for TB, Primary Active Disease, Active TB, Symptomatic – Clinical, Stage 4, Breakdown of Immune Control (Active TB), Definite TB | 36 | 97% |
| **State VI** | High severity of disease due to clinical features, radiology or bacteriology | **Cavitary/Disseminated, Active TB Disease - Symptomatic Clinical Smear Positive / Extensive Disease, Dissemination disease,** Active, Severe, Symptomatic - Culture Positive - Smear Positive, | 11 | 30% |
| **State VII** | Previous History of TB | **Asymptomatic Past TB, No Disease (Past TB), Treated / Recovered,** Treated, Old Disease, Relapse, Reinfection | 5 | 14% |

*Three most frequently used terms for each state are highlighted in bold. Abbreviated terms have* *been included as described in the article text, tables or diagrams. Tuberculosis (TB),* *Mycobacterium tuberculosis (Mtb), Latent Tuberculosis Infection (LTBI).*

**Table 3:** TB states described by articles included in the review

| Reference | State<br>0 | State<br>I | State<br>II | State<br>III | State<br>IV | State<br>V | State<br>VI | State<br>VII |
| --- | --- | --- | --- | --- | --- | --- | --- | --- |
| Barry 3rd et al.<br>(2009) | X | X | X | X |  | X |  |  |
| Young et al.<br>(2009) | X | X | X | X |  | X |  |  |
| Chao and Rubin<br>(2010) |  | X | X | X |  | X |  |  |
| Achkar and Jenny-Avital<br>(2011) |  |  | X | X |  | X | X | X |
| Gideon and Flynn<br>(2011) | X | X | X |  | X | X | X |  |
| Kunnath-Velayudhan et<br>al.<br>(2011) |  |  | X |  | X | X | X | X |
| Lawn et al.<br>(2011) | X | X | X |  | X | X |  |  |
| Sridhar et al.<br>(2011) | X | X | X | X |  | X |  |  |
| Esmail et al.<br>(2012) | X | X | X |  | X | X | X |  |
| Joel D. Ernst<br>(2012) | X |  | X | X |  |  | X |  |
| Delogu et al.<br>(2013) | X |  | X |  |  | X |  |  |
| Dowdy et al.<br>(2013) |  |  | X |  | X | X |  | X |
| Delogu and Goletti<br>(2014) | X | X | X |  |  | X |  |  |
| Esmail et al.<br>(2014) |  | X | X | X | X | X |  |  |
| Nunes-Alves et al.<br>(2014) |  | X | X |  |  | X |  |  |
| Salgame et al.<br>(2015) | X | X | X | X | X | X | X |  |
| Pai and Behr<br>(2016) | X | X | X |  | X | X | X |  |
| Pai et al.<br>(2016) | X | X | X |  | X | X |  |  |
| Petrucchioli et al.<br>(2016) | X | X | X | X | X | X |  |  |
| Cadena et al.<br>(2017) |  | X | X | X |  | X |  |  |
| Drain et al.<br>(2018) | X | X | X | X | X | X |  |  |
| Kik et al.<br>(2018) |  | X | X | X | X | X |  |  |
| Lin et al.<br>(2018) | X | X |  |  | X | X | X |  |
| Sousaa and Saraiva<br>(2018) | X | X | X | X |  | X |  | X |
| Houben et al.<br>(2019) |  | X | X | X | X | X |  |  |
| Lewinsohn and<br>Lewinsohn<br>(2019) | X | X | X |  | X | X |  |  |
| McHenry et al.<br>(2020) | X | X | X |  |  | X |  |  |
| Boom et al.<br>(2021) | X | X | X | X |  | X | X |  |
| Kendall et al.<br>(2021) | X | X | X | X | X | X |  | X |
| Migliori et al.<br>(2021) | X | X | X |  | X | X |  |  |
| Alebouyeh et al.<br>(2022) |  | X |  | X | X | X |  |  |
| Chandra et al.<br>(2022) | X | X | X |  |  | X | X |  |
| Esmail et al.<br>(2022) |  |  | X | X | X | X | X |  |
| Houben et al.<br>(2022) |  |  |  | X | X | X |  |  |
| Scriba et al.<br>(2022) | X |  | X |  | X | X |  |  |
| Yoon et al.<br>(2022) |  | X | X | X | X | X |  |  |
| Sossen et al.<br>(2023) |  |  | X | X | X | X |  |  |

Within the eight states, we identified an additional 27 unique sub-states (Supplementary Table 2). These were variations or minor differences within the conceptual themes that we had used to categorize. For States 0-I, conceptual variations included an acquired immune response that was detectable versus no longer detectable and elimination of infection through administration of preventative treatment. Conceptual sub-states for States II-IV differed in diagnostic results such as detection of granulomas on imaging. State V had the largest number of sub-states with articles describing variations in detection on imaging, bacteriological positivity and care-seeking behavior.

### Diagnostic criteria of TB states

Diagnostic criteria for TB states were described on the basis of tuberculin skin testing (TST), interferon-gamma release assay (IGRA), computerized tomography (CT), positron emission tomography combined with computerized tomography (PET-CT), X-ray, sputum culture, smear and Xpert and symptomatology. Articles varied in the use of diagnostic tests to differentiate states. Due to the variation in the descriptions of diagnostic tests described, we coded these into 5 “diagnostic domains”. These domains were: 1) Immune sensitization (including IGRA, TST); 2) High-resolution imaging (CT and PET-CT); 3) Chest X-ray; 4) bacteriology (including all sputum based investigations) and 5) symptomatology. The proportion of articles describing each diagnostic domain as either positive or positive/negative are displayed in Figure 2. Diagnostic definitions or symptomatology were described in 27 (67% of total) articles either part of diagrams, tables or within texts. Diagnostics were frequently not described for all states and not every diagnostic domain was included in articles. Articles where the outcome of a diagnostic test or symptom screen was not explicitly stated were categorized as missing and not included in the numerator. Articles where some diagnostic information was provided but had missing information for a particular diagnostic for a state have been retained in the denominator.

**Figure 2.**
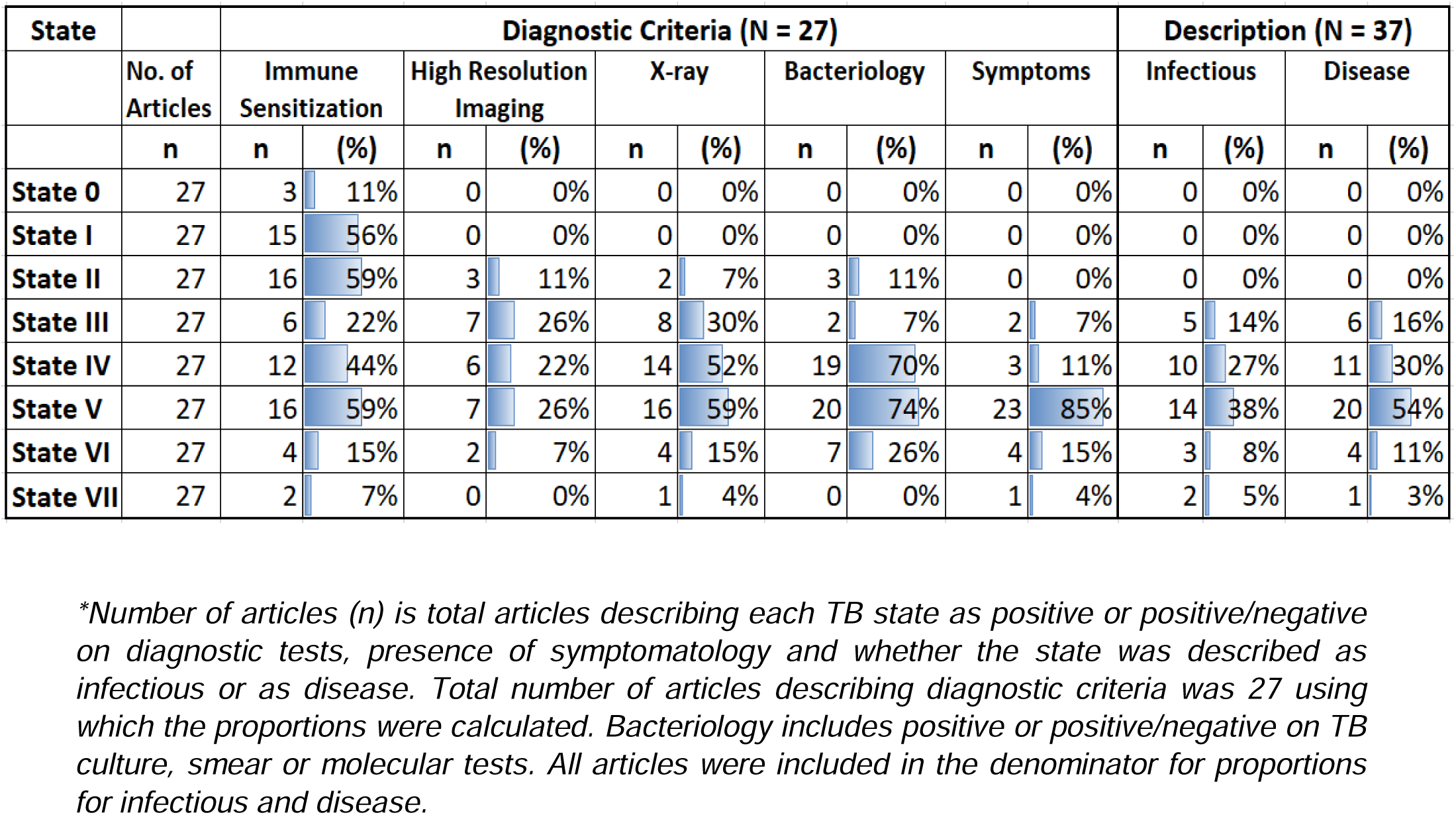
Diagnostic criteria and description of TB states

**State 0** was described as negative on all diagnostic tests with the exception of 3 (n=3/27; 11%) articles describing positive/negative results on immune sensitivity tests. Individuals in **State I** were most frequently described as having a positive diagnostic result to immune sensitization tests (n=15/27; 56%) and negative to all other tests. Limited diagnostic differentiation was described for **State II** with articles most frequently describing positive immune sensitivity tests (n=16/27; 59%), followed by positive CT/PET-CT (n=3/27; 11%)). **State III** was most frequently described to be positive on X-ray (n=8/27; 30%) and PET-CT/CT (n=7/27; 26%). **State IV** was frequently described as having positive bacteriologically (n=19/27; 70%) with remaining articles describing this state as subclinical only, without providing explicit diagnostic results. **State V** was most commonly described as having positive symptoms (n=23/27; 85%) with the remaining articles not explicitly stating whether this state would yield positive symptom screen results. **State VI** was most frequently associated with positive bacteriology (n=7/27; 26%) and symptoms (n=4/27; 15%). **State VII** was most frequently associated with positive immune sensitivity (n=2/27; 7%).

### Infectiousness and disease threshold

To assess the epidemiological significance of each disease state, articles were reviewed for the terms *“infectious”* or *“transmissible.”* To understand at which state articles considered TB disease to begin (disease threshold), the term *“disease”* was reviewed. These terms were included in the terminology, narrative descriptions or as part of conceptual diagrams for disease states. No article described States 0, I or II as infectious or as disease. States IV and V were most frequently described as infectious (n=10/37; 27% and n=14/37; 38% respectively) and as disease (n=11/37; 30% and n=20/37; 54% respectively).

## Discussion

This is the first review to systematically map and synthesize evidence from published literature for additional TB states beyond the prevailing latent and active paradigm. We identified 37 articles relevant to our inclusion criteria, a third of which were published within the past three years, suggesting increasing interest and recognition of additional states for TB. There are several key themes that emerged from our review.

There was significant variation in the frequency of TB states and in their conceptual definitions, highlighting an overall lack of consensus within literature. Articles varied in their focus and emphasis, for example, TB pathogenesis and bacteriology, immunology, clinical or epidemiological relevance, diagnostic biomarkers and imaging were utilized as different perspectives for the basis of TB states. These perspectives framed the conceptual approach towards TB states and subsequent inclusion or exclusion of particular states. The choice of which states to include may also have been driven by prevailing knowledge gaps and research priorities at the time of the article publication. Earlier articles (2009 – 2015/16) were broadly more focused on TB immunology and host mechanisms of sterilization of *Mtb* and have therefore described states with innate and adaptive clearance. More recent articles tended to shift focus towards epidemiological considerations including detection and transmission implications of asymptomatic disease. As a result, states describing subclinical disease were more common. While we aimed to develop a parsimonious set of states grouped around common concepts, there were variations within these themes that we described as *sub-states*. Certain articles chose to further elaborate upon sub-states and this again reflected the focus of the article and the perspective of the authors.

The review also highlights a lack of consistency in the terminologies used to describe TB states. We observed that not only were different terms utilized to describe the same concept within the natural history of disease but similar terms were also utilized to describe different conceptual states. As examples, bacteriologically positive disease in the absence of symptoms was variously described as *subclinical, asymptomatic, incipient* and *bacteriologically positive*. Similarly, *latent TB* was utilized to describe elimination of infection with an identifiable acquired immune response, containment of infection, as well as breakdown of immune control. Such variation in nomenclature naturally leads to confusion in communication and conduct of scientific research and in public health policy. The understanding of a certain term in literature is subjective to the perspective of the authors and interpretation of the readers and these may not necessarily be the same.

Two-thirds of articles outlined diagnostic criteria. Similar to the conceptual states, articles varied in the choice of diagnostic tests to differentiate between states. The lack of a standardized diagnostic criteria to differentiate between TB states may lead to further challenges in describing TB states. We chose to group diagnostic criteria within five domains and these may prove useful in developing diagnostic definitions for proposed conceptual states of TB. Further considerations include achieving consensus on defining the domains themselves. Bacteriology may be defined through culture, molecular tests, smear, as well as type of sampling, such as through sputum, aerosols or through use of induced and invasive methods. Similarly, symptomatology can include those who are truly asymptomatic, those who test negative on a symptom-screen test or those who are symptomatic but do not seek clinical care.

From a public health and individual perspective respectively, the point at which TB is considered infectious to others or as causing disease (and hence harm to the individual) is important. It is notable therefore that the majority of articles did not directly address this concept. Infectiousness of TB states was described by less than half of articles. No articles considered State 0, I or II to be infectious, with this proportion steadily increasing from State III (infection not controlled by immune system) to State IV (bacteriologically positive in the absence of symptoms) and State V (clinically apparent symptomatic TB). Surprisingly, no articles explicitly describe where they believed the disease threshold to begin and this could only be inferred from terminology used in the text and in diagrams. The number of articles using the term disease again increased from State III through to State V.

While it is clear that there is an increasing recognition of the TB spectrum in recent literature, the various states outlined by the included articles are not all new. As our historical review of the NTA proceedings highlights, TB in the past has been considered as having several stages, with staging systems reflecting the public health needs of the time. Several of these early disease states that have been highlighted in the contemporary literature we reviewed were also described in the pre-antibiotic era (Box 1). In the absence of effective treatments, it was recognized that screening, early-identification and prompt isolation of individuals with TB in sanatoria may help control the disease. Development of potent chemotherapy for TB, coupled with increased commitments for primary healthcare, shifted the focus away from mass X-Ray based case finding towards passive approaches towards case-finding and treatment (12). This cemented the binary conceptualization of TB, as individuals were either considered to be harboring “latent” infection or presenting to healthcare facilities with clinically apparent, “active” disease. It is possible that the reemergence of early TB disease states in literature over the past decade has coincided with increased investments for active case-finding for TB, such as through community-wide screening and use of mobile X-rays. In addition, improved understanding of TB pathophysiology as well as advances in development of biomarkers and imaging may have stimulated further interest in earlier disease states.

Our review underscores the need to develop a clear consensus on conceptual definitions, terminologies and diagnostic criteria for TB states. This consensus should involve a broad range of stakeholders so that agreed definitions and terminologies will be acceptable from research, clinical, public health and policy perspectives, in addition to being acceptable to individuals with TB. Standardized diagnostic definitions that can be consistently applied across different settings will require careful consideration and are an important area for further research.

TB has a complex pathophysiology and as we have shown it is possible to describe many states, hence the emergence of the commonly used phrase of “spectrum”. The binary representation of TB is clearly insufficient, however, incorporating any additional states must have a purpose and relevance to public health or to people with TB and should also facilitate the goal of global TB elimination. Parsimony and simplicity should remain key guiding principles for the development of this framework with clear language, diagnostic definitions and disease thresholds. This will be of critical importance in communication to researchers, care providers, people with TB and to the general public. Any framework should be regularly reviewed and adapted as our knowledge improves along with advances in diagnostics and treatment.

### Limitations

Our search strategy was restricted to review articles in English and it is likely that additional TB states or terminology may have been described in other types of articles and languages. However, since the aim of the scoping review was to provide an overview rather than to comprehensively document all described states, this restriction was considered appropriate.

As with other types of thematic analyses, the categorization and coding for TB states is to a degree subjective to the authors’ interpretation and views. This is particularly relevant for states where conceptual diagrams were not provided and categorization had to be inferred from the article text. Our goal was to categorize states around a minimum number of common concepts, not all variations described in the articles could be adequately captured within one of these categories. We attempted to resolve this by including sub-states where such distinctions and variations were categorized. Biases in coding were minimized by coding through two reviewers and resolution by a third reviewer in cases of disagreement. All study authors reviewed the final set of states and their diverse expertise and backgrounds helped further minimize bias.

Articles varied in the diagnostic tests and criteria utilized to classify diseases states depending on the nature of the article and background of the authors. This led to a number of articles with missing information on diagnostics, symptomatology, disease description and transmission potential for those disease states. To remove potential for biases, a diagnostic test or criteria was not included in the positive and positive/negative category unless explicitly stated in the article. The ratios of positivity on diagnostic criteria are therefore lower than would be expected had each article described them and should therefore be interpreted with caution and not as precise measurements of the authors’ opinions.

## Conclusion

Early states of TB beyond latent and active disease have been increasingly described in literature over the past decade. However, the conceptual definitions and diagnostic criteria for these states and the terminology used to describe them is highly inconsistent. Consensus on a framework for describing early TB disease states is therefore required. This will help standardize how early TB disease is reported in literature, inform clinical and public health management as well as guide research for TB elimination.

## Data Availability

All data produced in the present study are available upon reasonable request to the authors

## Acknowledgements

We would to thank Heather Chesters, Deputy Librarian, UCL Great Ormond Street Institute of Child Health Library, UCL Library Services for support with the literature search.

