## Supplementary File for "Beyond latent and active – a scoping review of conceptual frameworks and diagnostic criteria for tuberculosis"

### Supplementary Table 1: Search terms utilized in Ovid in scoping review

| 1 | *Mycobacterium tuberculosis/ or *Latent Tuberculosis/ or *Tuberculosis, Pulmonary/ or *Tuberculosis/ |
| --- | --- |
| 2 | spectrum.mp. |
| 3 | framework.mp. |
| 4 | concept*.mp. |
| 5 | paradigm.mp. |
| 6 | stage.mp. |
| 7 | state.mp. |
| 8 | 2 or 3 or 4 or 5 or 6 or 7 |
| 9 | 1 and 8 |
| 10 | "Review Literature as Topic"/ or "Systematic Review"/ or "Review"/ |
| 11 | 9 and 10 |

### Supplementary Table 2: Mycobacterium *tuberculosis* sub-states identified from articles included in scoping review

| **State / Sub-State** | **Concept** | **n** | **(%)** |
| --- | --- | --- | --- |
| **State 0** | **Infection has been eliminated by innate immune response** | **23** | **62%** |
| State 0A | Innate resisters (individuals naturally resistant to acquiring infection) | 8 | 22% |
| **State I** | **Infection has been eliminated by acquired immune response** | **28** | **76%** |
| State IA | Infection eliminated through specific immune mechanism that is detectable | 15 | 41% |
| State IAa | Infection eliminated through specific immune mechanism that is no longer detectable (reversion) | 12 | 32% |
| State IAb | Infection eliminated through specific immune mechanism >2 years | 1 | 3% |
| State IB | Infection eliminated through TPT | 3 | 8% |
| State IBb | Infection eliminated through TPT (INH) | 5 | 14% |
| State IBb | Infection eliminated through TPT (RIF) | 5 | 14% |
| **State II** | **Infection is not eliminated but is controlled by the immune system** | **34** | **92%** |
| State IIA | Infection is not eliminated, controlled in granuloma (not detectable, replicating bacteria) | 13 | 35% |
| State IIB | Infection is not eliminated, controlled in granuloma (detectable) | 3 | 8% |
| State IIC | Infection is not eliminated, controlled in granuloma (not detectable, non-replicating bacteria) | 15 | 41% |
| **State III** | **Infection is not controlled by the immune system** | **21** | **57%** |
| State IIIA | Infection is not eliminated, not controlled in granuloma (detectable) | 8 | 22% |
| **State IV** | **Individual is bacteriologically positive but has no symptoms or clinical features** | **23** | **62%** |
| State IVA | Detectable bacteria but no symptoms (additional sampling) | 2 | 5% |
| State IVB | Detectable bacteria but no symptoms (Bac +ve, CXR -ve) | 13 | 35% |
| State IVBa | Detectable bacteria but no symptoms (Bac +ve, CXR +ve) | 14 | 38% |
| **State V** | **Individual has signs or symptoms associated with TB** | **36** | **97%** |
| State VA | Symptomatic or Clinical Disease (Bac -ve) | 6 | 16% |
| State Aa | Symptomatic or Clinical Disease (Bac -ve, CXR -ve) | 1 | 3% |
| State VAb | Symptomatic or Clinical Disease (Bac -ve, CXR +ve) | 4 | 11% |
| State VB | Symptomatic or Clinical Disease (Bac +ve) | 23 | 62% |
| State VBa | Symptomatic or Clinical Disease (Bac +ve, CXR -ve) | 1 | 3% |
| State VBb | Symptomatic or Clinical Disease (Bac +ve, CXR +ve) | 16 | 43% |
| State VBc | Symptomatic or Clinical Disease (Bac +ve, smear -ve) | 4 | 11% |
| State VC | Symptomatic but not care seeking | 3 | 8% |
| **State VI** | **High severity of disease due to clinical features, radiology or bacteriology** | **11** | **30%** |
| State VIA | Disseminated disease | 8 | 22% |
| State VIB | Cavitary Disease | 10 | 27% |
| **State VII** | **Previous History of TB** | **5** | **14%** |
| State VIIA | Repeat Diagnosis after previously cured of TB (recurrent) | 2 | 5% |
| State VIIB | Cured after TB Diagnosis | 5 | 14% |
| State VIIC | Death | 2 | 5% |

*TB states concepts that included variations within the 8 major themes identified were categorized as sub-states. Articles described either 1) the state only; 2) the state and sub-state; or 3) the sub-state only. Since sub-states were not described in several articles, their summation does not equal the number for states. In cases where only a sub-state was described, it was included in the number for the state as well. TB Preventative Treatment (TPT). Isoniazid (INH). Rifampicin (Rif). Bacteriologically positive (Bac +ve). Bacteriologically negative (Bac -ve).Tuberculosis (TB).*
